# Model of a Testing-and-Quarantine Strategy to Slow-Down the COVID-19 Outbreak in Guadeloupe

**DOI:** 10.1101/2020.05.01.20088138

**Authors:** Meriem Allali, Patrick Portecop, Michel Carlès, Dominique Gibert

## Abstract

Using a stochastic epidemic model explicitly considering the entire population of Guadeloupe (1), we explore the domain of solutions presenting an efficient slowing down of the COVID-19 epidemic spread during the post-containment period. The considered model parameters are the basic reproduction number *R*_0_ to simulate the effects of social distancing, the time delay *δT_q_* elapsed between the detection of a symptomatic person and her/his placement in quarantine to suppress her/his contagiousness, and the number *N_a_* of asymptomatic people tested positively and isolated. We show that acceptable solutions are obtained for a wide range of parameter values. Thanks to a good control of the initial epidemic spread resulting from an early containment and efficient communication by the sanitary and administrative authorities, the present situation corresponds to a pre-epidemic state. The most safe solutions are a combinations of social distancing, numerous testing to perform a systematic isolation of symptomatic patients and guided detection of asymptomatic people in the entourage of localised symptomatic patients.

## Introduction

For the time being, the COVID-19 disease caused by SARS-CoV2 continues is rapid worldwide spreading and, according to the World Health Organisation (Wor, 2020c), as of May 1, 2020, more than 3 millions of people have been infected causing the death of about 230,000 persons (2). This has led States to take drastic measures of containment to reduce and hopefully stop the propagation of SARS-CoV2 across the population (3,4). However, containment may not last beyond two or three months without causing collateral devastating psychological (4–6), social and economical effects (7). For instance, after a full month of containment, a growing part of the French population is already asking for a partial release of the containment conditions and is concerned with the post-containment procedures (8).

Several countries seem to have passed the maximum of the epidemic crisis and are now preparing their post-containment epoch. Such is the case for France where the daily number of new infected persons seems to begin to decrease slowly (9). On April 13, the President of the French Republic, Mr Macron, indicated that the end of strict containment period could hopefully start on May 11, 2020. However, it is widely accepted that any uncontrolled ending of containment would undoubtedly result in an exponential restart of the COVID-19 infection (10). The main reason is that the number of noninfected represents a large fraction of the population, giving to the virus a way to again propagate from only few infectious people. It is of a primary importance to explore and document the positive and negative effects of the different measures that could be envisioned to, at least partly, release the containment for a larger as possible part of the population.

Whatever the means involved to control the end of the containment stage, they all have the objective to reduce the basic reproduction number, *R*_0_, to an as small as possible value below 1 in order to suppress any exponential divergence of the number of infected (and also infectious) persons. In the case of COVID-19, keeping *R*_0_ at or slightly above 1 would reduce the instantaneous number of infected people and, subsequently, the instantaneous number of severely affected patients needing intensive cares. This procedure maintains the load of intensive care units below their saturation and guaranties that every patient will benefit of adequate cares. Ultimately, the whole population will have been contaminated and protected against a novel infection. However, because of the infection fatality rate of COVID-19, about 1 – 2% of the population will decease (11, 12). In France, this would represent the death of approximately 1 million persons. It is preferable to reduce *R*_0_ below 1 in order to stop the spread as quickly as possible and make the absolute number of infected persons as small as possible (3).

The present study is a sequel of an article (1) where we propose a predictive model of COVID-19 spreading in Guadeloupe. Presently, this model continues to accurately predict the new data daily released by the sanitary authorities. In the present paper, we evaluate the efficiency of testing and isolating people affected by COVID-19. We also consider the global effects of social distancing through a control of the basic reproduction number *R*_0_ (3). By using a slightly modified version of the model of Allali et al. we give estimates of the efficiency of the testing-isolating-distancing combination of measures to produce a progressive shutdown of the COVID-19 spread in Guadeloupe.

## Description of the model

The stochastic model proposed by Allali et al. has been adapted to introduce perturbing operations in the normal epidemic spread process described in our previous article. These perturbing operations mimic sanitary procedures that modify the contamination chain and their role is to lower the global basic reproduction number *R*_0_. We hereafter describe only the changes applied to the model. The reader is referred to Allali et al. for a detailed description of the initial model.

The main control parameter to consider is the *R*_0_ reproduction number. However, this number encompasses the various ways by which the contamination by SARS-CoV2 proceeds and, to reduce *R*_0_, it is necessary to account for the different step-process over which one may have some action. Following Grassly and Fraser, Ferretti et al. decompose the basic reproduction number of the COVID-19 spread as a sum of four contributions representing asymptomatic transmission *(R_a_)*, pre-symptomatic transmission *(R_p_)*, symptomatic transmission (*R_s_*), and environmental transmission (*R_e_*):

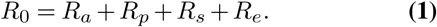

Relying upon a review of clinical and epidemiological data, Ferretti et al. use: *R_a_ =* 0.1, *R_p_ =* 0.9, *R_s_ =* 0.8 and *R_e_ =* 0.2 whose sum gives *R*_0_ = 2.0. These partial reproduction numbers are themselves the integrals of non-uniform infectiousness probability functions *ξ* (noted *β* by Ferretti et al.). Because of their relative high values, both *R_p_* and *R_s_* are the main quantities to reduce in a spread slow-down protocol. To reach this objective, procedures aimed at stopping the contagiousness of a fraction of infected patients are considered. Owing to the fact that testing everybody everyday is impossible, even for the 400,000 inhabitants of Guadeloupe, we consider that the new symptomatic patients are good entry points. In the present study, we assume that any new symptomatic person is identified, tested and placed in quarantine in a time delay *δt_q_* after the appearance of the symptoms. By this way, *R_s_* is diminished by reducing the duration of contagiousness of symptomatic patients.

A reduction of *R_p_* may be obtained by detecting asymptomatic patients in the entourage of every detected symptomatic patient. Such a guided detection procedure is efficient because asymptomatic persons are more likely to be found near symptomatic patients. Testings are consequently more efficiently performed. Because the detection procedure is causal, it is impossible to be sure that all detected asymptomatic people are future pre-symptomatic ones in the sense of Ferretti et al.. In the present study, we make no distinction between asymptomatic and pre-symptomatic because such a distinction can only be made *a posteriori*. The average number, *δN_a_*, of detected asymptomatic people is a parameter whose threshold value necessary to obtain *R*_0_ < 1 has to be determined by simulation.

Another action, not considered in our model, is to perform testing on people having high social contacts like, for instance, nurses, teachers, cooks, etc.. and who could have a high personal *R*_0_.

Figure 1 shows the flowchart of all possible evolution schemes for the “asymptomatic” and “symptomatic” classes of the model. At time *T*_1_, some susceptible people are infected and become asymptomatic patients *A*_1_*,A*_2_*,A*_3_*,A*_4_.

**Fig. 1.**
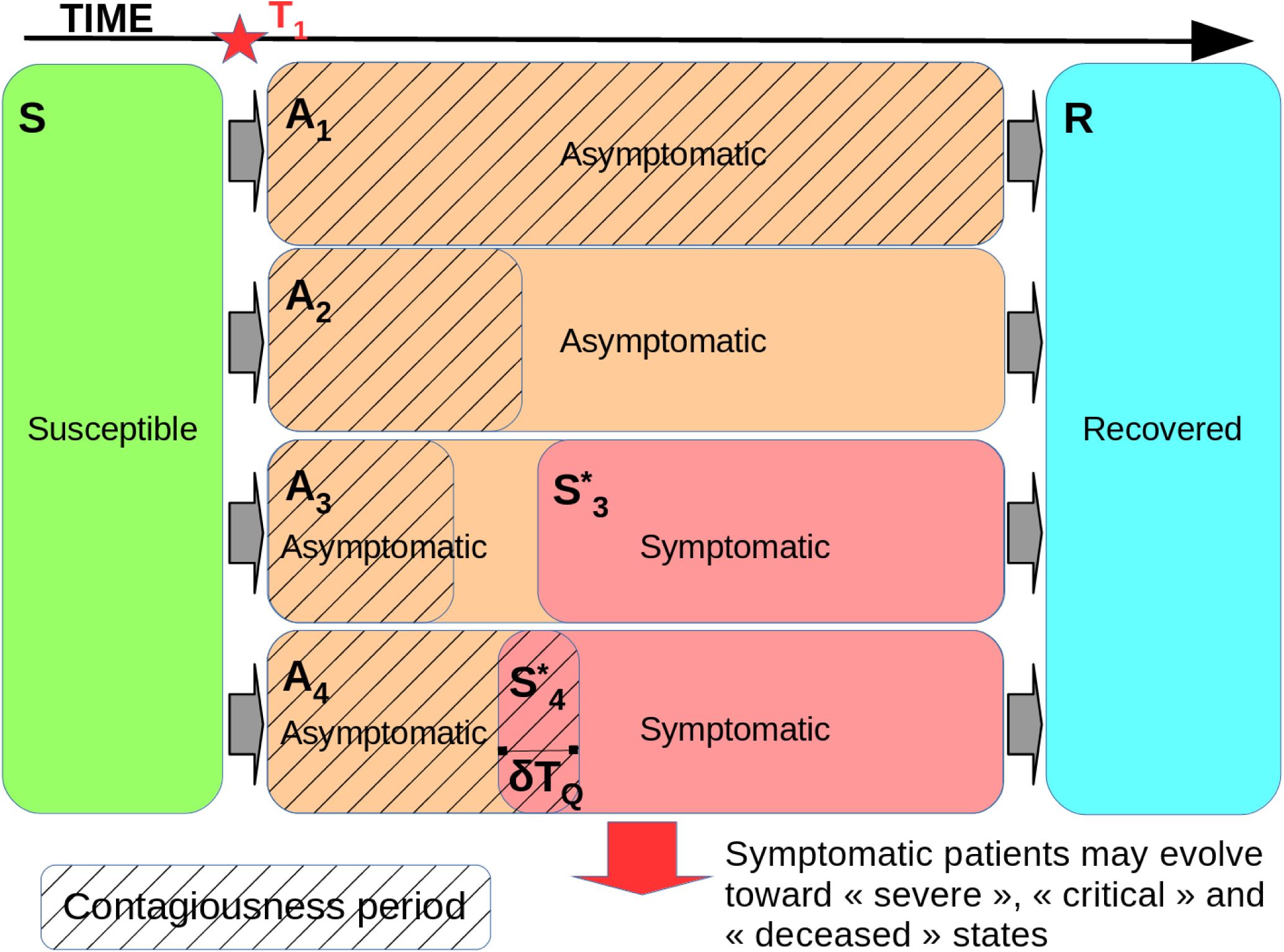
Flowchart of the possible evolution schemes for the “asymptomatic” and “symptomatic” classes of the model. At time *T*_1_, 4 susceptible persons are infected. At the beginning, all four patients are asymptomatic (orange areas) and contagious (hatched areas). When becoming symptomatic (red areas), patient *S*_4_ is identified, tested and isolated. The *δT_Q_* parameter represents the time-delay elapsed between the apparitions of the symptoms and the moment when the patient is placed in quarantine to suppress her/his contagiousness. This marks the end of her/his contagiousness period. Asymptomatic patients (e.g. *A*3 and *A*2 located in the entourage of *S*_4_ are tested and isolated. Patient *A*_1_ remains asymptomatic and undetected all along her/his illness period. In the model, patients *S*_3_ and *S*_4_ may evolve to other states: “severe”, “critical” and “deceased” (see (1) for details).

Then, some time later, patient *A_4_* becomes symptomatic *(S_4_)* and is detected and tested within a time delay *δt_q_* after the apparition of the first symptoms. Once detected, patient *S*_4_ is placed in quarantine and is no more contagious. This results in a decrease of *R_s_*. At the same time, tests are performed on people in the entourage of *S*_4_ and asymptomatic persons *A*_2_ and *A*_3_ are detected and also placed in quarantine, producing a decrease of *R_a_*. In the flowchart of Figure 1, the asymptomatic patient *A*_1_ remains undetected all during the contagiousness period and is able to contaminate other persons. Both *S*_3_ and *S*_4_ patients may either recover or evolve toward “severe”, “critical” and “deceased” states (Fig. 2).

**Fig. 2.**
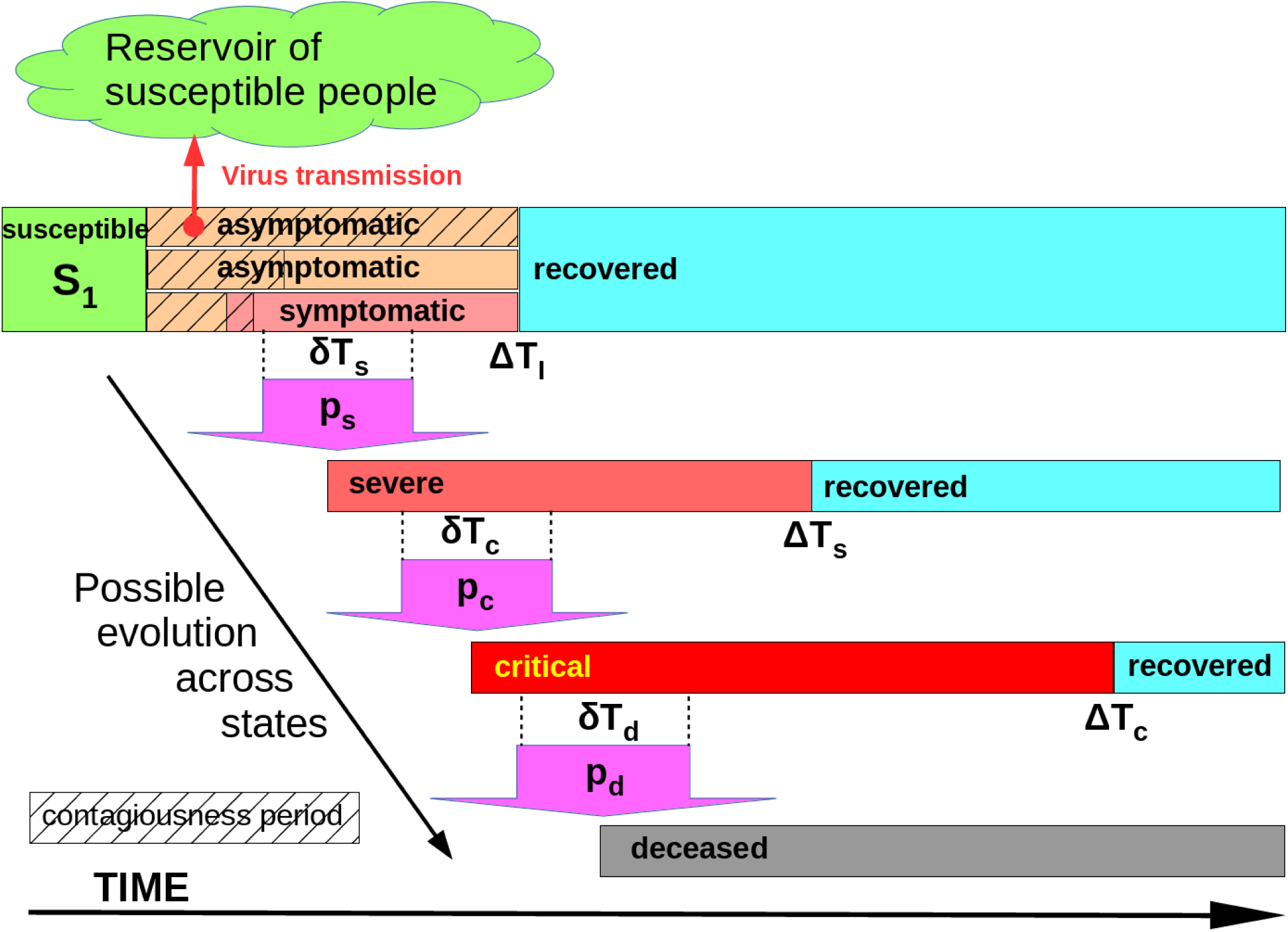
Flowchart of the stochastic modelling procedure for the model proposed by Allali et al. including the detailed asymptomatic/symptomatic sequence of Figure 1. In this modified version of the model, a symptomatic patient may evolve toward the “severe” state, the “critical state and the “deceased” state with respective probability *p_s_*, *p_c_* and *p_d_*. The transition from one state to the next may only occur within time windows noted *δT_s_, δT_c_* and *δT_d_*. The recovery time represents the duration after which a patient in a given state is declared “recovered”, they are noted Δ*T_s_*, Δ*T_s_* and Δ*T_c_* respectively for the states “symptomatic and asymptomatic”, “severe” and “critical”.

To implement the evolution scheme of Figure 1, the stochastic model proposed by Allali et al. has been modified to simulate the very beginning of the period of illness, when newly infected patients may switch from asymptomatic to symptomatic. By this way, we are able to model the date of appearance of symptoms and decide to apply prescribed testing and containment procedures. The class of infected people “minor” in the previous model has been split into 2 classes, namely: “asymptomatic” and “symptomatic” people. As said above, the causal nature of our model makes impossible to distinguish “asymptomatic” from “pre-symptomatic” persons as considered by Ferretti et al.. In the present study, these two classes have been merged into a single one called “asymptomatic”.

The contagiousness function *ξ_c_* corresponding to the “pre-symptomatic” class is also assigned to our “asymptomatic” class. This makes the model more pessimistic in order to make the conclusions more constraining than needed in reality. In the present study, we set *ξ_c_* as a gamma distribution with shape and scale parameters respectively equal to 5 and 1.

Following in Linton et al., we represent the switching function ζ*_a/s_* by a lognormal probability function with mean μ_ζ_ and standard deviation to randomly draw the dates of switching from “asymptomatic” to “symptomatic”. In the present article, we use the right-truncated values of *μζ* = 5.6 days and *σ_z_eta* = 3.9 days proposed by Linton et al..

Figure 2 shown the general flowchart of the stochastic model with the new asymptomatic/symptomatic sequence included. The other states are identical to those of the model of Allali et al.. The meaning of the different parameters is given in the Figure legend and all input parameters are listed in Table 1.

**Table 1.** Model input parameters. The three first top parameters in the Table (*R*_0_, *δT_Q_*, *N_a_*) have changing values in the different simulations. Other parameters remain unchanged for all simulations.

| Parameter name | Symbol | Value | Comment |
| --- | --- | --- | --- |
| Basic reproduction number | $R_0$ | 1.5, 2, 3, 4 | Time-varying parameter through the contagiousness function $\xi_c$. |
| Detection delay | $\delta T_Q$ | 1, 2, 3, 4 | This parameter is a time difference defined as the delay (in days) between the apparition of symptoms in a patient and the moment when this patient is placed in quarantine. This represents the time delay taken by the sanitary authorities to tackle stop the contagiousness of a symptomatic patient. |
| Number of detected asymptomatic | $N_a$ | 0 to 4 | Number of asymptomatic people detected in the entourage of each detected symptomatic patient. |
| Number of initial asymptomatic | $Z_I$ | 100 | This is a conservative value for $Z_I$, and the model fitting to the data available up to now (Fig. 3) indicates that a smaller value for $Z_I$ may be expected. The results obtained with $Z_I = 100$ may be proportionally multiplied or divided to account for different values of $Z_I$. |
| Switching function (lognormal) | $\zeta_{a/s}$ | $\mu_\zeta = 5.6$ (days)<br>$\sigma_\zeta = 3.9$ (days) | This probability function give the switching time dependence of the contagiousness of asymptomatic, pre-symptomatic and symptomatic people. $\zeta_{a/s}$ is taken as a lognormal distribution with mean and standard deviation given by Ferretti et al.. |
| Contagiousness function (gamma) | $\xi_c$ | $\alpha_\xi = 5$ (shape)<br>$\beta_\xi = 1$ (scale) | Represents the time dependence of the contagiousness of asymptomatic, pre-symptomatic and symptomatic people. $\xi_c$ is taken as a gamma distribution (15). In the model, the mean and standard deviation are respectively equal to 5 and 2.2 days. |
| Fraction of asymptomatic people | $\alpha_a$ | 0.20 | $\alpha_a$ represents the fraction of asymptomatic with respect to symptomatic patients. $\alpha_a = 0.2$ corresponds to 20% of asymptomatic and 80% of symptomatic. The value taken in this study is given by Mizumoto et al.. |
| Recovery time of asymptomatic and symptomatic | $\Delta T_s$ | 14 days | Patients who remained in a given state during the corresponding recovery time is automatically switched to the "removed" state. |
| Recovery time of critical | $\Delta T_c$ | 21 days | |
| Recovery time of severe | $\Delta T_I$ | 14 days | |
| Switching period from symptomatic to severe | $\delta T_s$ | 4-10 days | Time window during which a patient may switch to the next evolution state. For example, a "severe" patient may switch to "critical" from day 0 to day 4. After day 4, the patient is supposed medically stabilised. The switching date is randomly drawn in the time window. |
| Switching period from severe to critical | $\delta T_c$ | 0-4 days | |
| Switching period from critical to deceased | $\delta T_d$ | 2-4 days | |
| Switching probability from symptomatic to severe | $p_s$ | 0.14 | Probability to switch to the newt state during the switching period. |
| Switching probability from severe to critical | $p_c$ | 0.25 | |
| Switching probability from critical to deceased | $p_d$ | 0.35 | |

## Data

The data used in the present study, are daily communicated by the University Hospital to the local authorities, i.e. the Regional Health Agency (Agence Régionale de Santé in French). They correspond to the cumulative number of persons with COVID-19, the cumulative number of deceased patients and the number of patients presently in intensive care units. Detailed data for France are made available by Santé Publique France (9) (see also Alamo et al. for a review of open data repositories).

Both the cumulative number of deceased patients and the number of patients presently in intensive care units respectively correspond to ∑*N_d_* and *N_c_* in the model. The cumulative number of persons with COVID-19 could be something between ∑*N_I_* and ∑*N_s_*, depending on the screening procedure. In France, a majority of the persons tested for COVID-19 are patients with severe symptoms and admitted in specialised COVID-19 units. Such is the case in Guadeloupe and, consequently, the cumulative number of persons with COVID-19 as announced by ARS correspond to the ∑*N_s_* of the model.

In the present study, we use the data going from March 13 2020 to April 21 2020 shown in Figure 3. The model shown on this Figure is an update of the model prediction model proposed by Allali et al.. The parameter values of the updated model are almost identical to those of the former model despite the fact that the contagiousness function ξ*_c_* is now nonuniform in time.

**Fig. 3.**
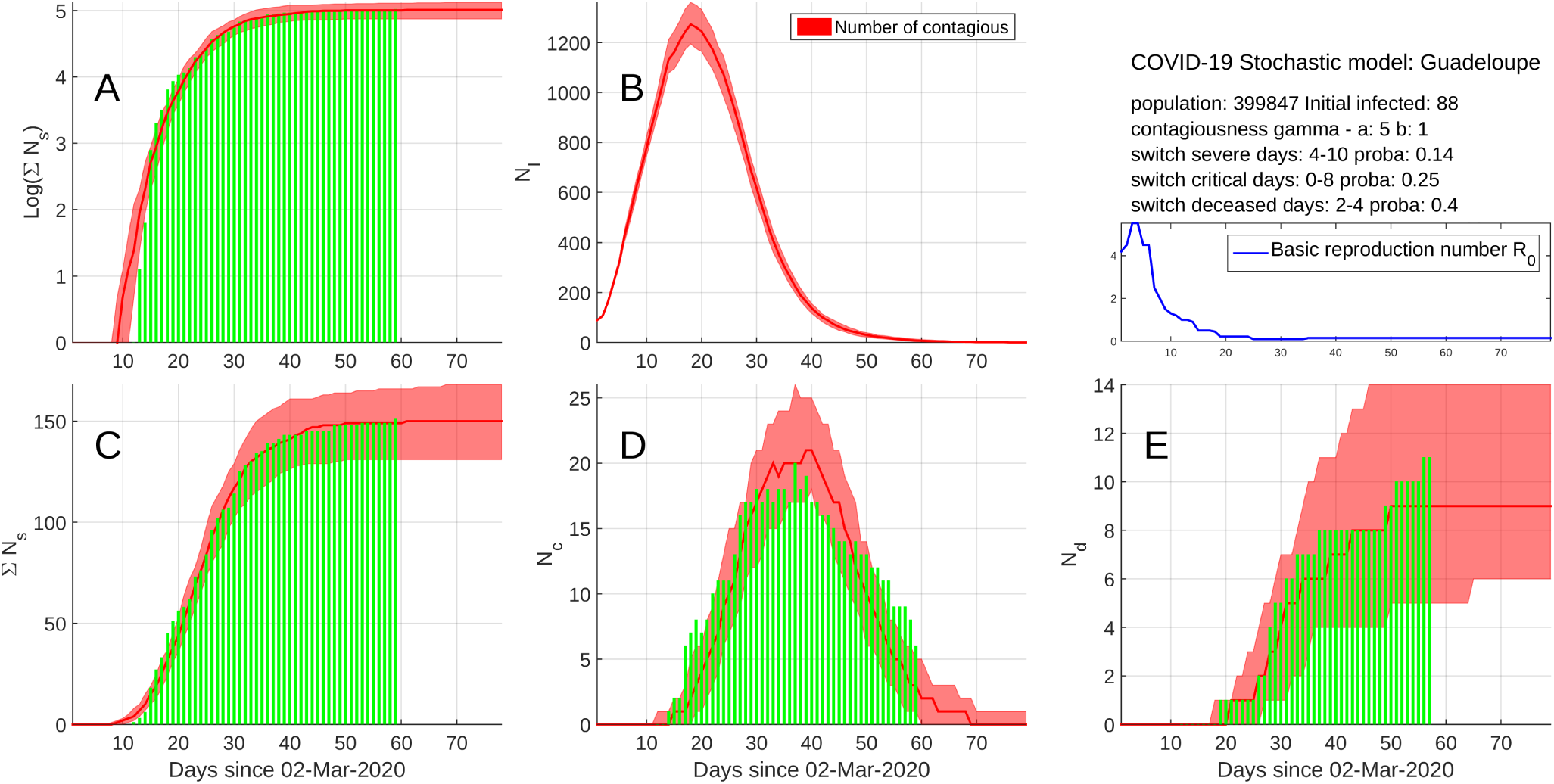
Model 1 results. A) semi-logarithmic (Natural logarithm) plot of the cumulative number ∑*N_s_* of severe cases. Green bars = data and red bars = model. B) Instantaneous number *N_I_* of infectious. C) same as (A) in linear axis. D) Instantaneous number *N_c_* of critical cases. Green bars = data and red bars = model. E) Cumulative number ∑*N_d_* of deceased patients. Green bars = data and red bars = model. The parameter values used in the model are shown in the upper-right part of the figure together with the time-variation of the basic reproductive number *R*_0_. The red rectangles represent the 80% confidence interval centred on the median.

## Results

The principal parameters that may represent the various measures taken to control the epidemic spread during the post-containment period are *r*_0_, δ*t_q_* and *N_a_* (Table 1). The basic reproduction number *R*_0_ represents the overall effect of social distancing and basic hygiene measures. The time delay *δt_q_* represents the duration between the onset of symptoms in a patient and the moment when this patient is identified, tested and isolated. The number *N_a_* is the average number of asymptomatic people that are supposed to be identified in the entourage of a detected symptomatic patient. We assume that these asymptomatic patients are isolated simultaneously with the symptomatic person.

We shall assume that all other parameters are kept unchanged for all simulations. The values of these parameters are either taken from literature (17) or determined from the fit with the data (Figure 3). The parameter values are given in Table 1. We performed numerous simulations for triplets *(R*_0_*,δT_Q_,N_a_)* in the ranges: *R*_0_ ∊ (1.5,2.0,3.0,4.0), δ*t_q_* ∊ (1,2,3,4) and 0 ≤ *N_a_ ≤* 4. Depending on the values chosen for the parameters, the model solutions fall into 3 classes:

**Diverging solutions**. An example of such a solution is shown in Figure 4 which corresponds to an uncontrolled situation where the exponential growth (Fig. 4A,F) of infected people continues until the entire population is affected (Fig. 4E). These solutions inevitably conduct to a very large number of critical (*∑N*_c_ ≈ 10500) and deceased patients *(∑N_d_* ≈ 2800, Fig. 4D). Because of the large instantaneous number *N_c_* of critical patients, intensive care units are overloaded and many patients may not receive adequate cares. This failure of sanitary facilities would induce additional deceased people, i.e. roughly the sum ∑*N_c_* + ∑*N_d_*.

**Fig. 4.**
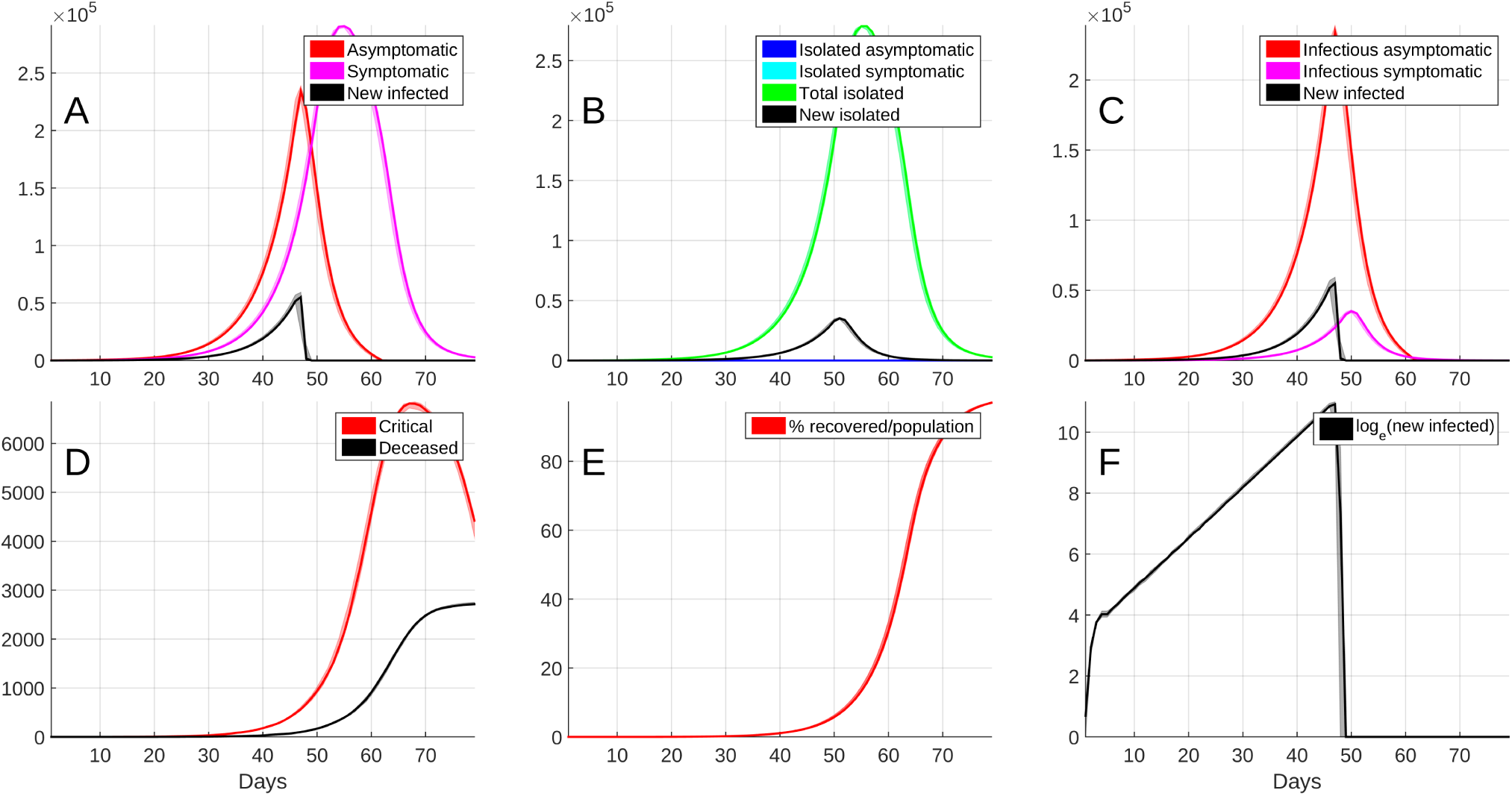
Example of diverging solution obtained for *R*_0_ = 4, δ*T_Q_* = 1 day and *N_a_* = 0. The curves represent the median of 40 runs of the model with the same initial conditions. A) Instantaneous number of asymptomatic *(N_a_*), symptomatic *(N_s_*) and newly infected *(N_I_*) people. B) Instantaneous number of isolated symptomatic and newly isolated patients. In this example, no asymptomatic persons are isolated and the total number of isolated people equals the number of isolated symptomatic. C) Instantaneous number of non-isolated (i.e. infectious) symptomatic and asymptomatic people. Also shown is the number of newly infected person (also shown in A). D) Instantaneous number of patients in critical state and cumulative number of deceased patients. E) Percentage of the population infected. F) Semi logarithmic plot of the number of newly infected people (also shown in A and C with linear axes). The origin of the time axis corresponds to the beginning of the post-containment period.

**Slowdown solutions**. These solutions correspond to controlled situations where the exponential spread is rapidly stopped and followed by a sharp damping of the number of new infected people (Fig. 5A). The short exponential growing observed at the beginning of the spread occurs during the delay when no symptomatic patients are declared and isolated. In the model, this delay corresponds to the mean of the log-normal distribution (see Table 1). For this kind of solutions, the number of critical and deceased patients remains small (Fig. 5D) when compared with diverging solutions. The number of persons to isolated every day (black curve in Fig. 5B) may reach ≈ 200, and adequate logistical and testing facilities are necessary to treat this quantity of people. In case of failure of these facilities, the situation could rapidly become uncontrolled and diverging. The 50% envelopes of the model outputs obtained for the 40 runs (i.e. coloured areas) are narrow, indicating a stability of the stochastic process.

**Fig. 5.**
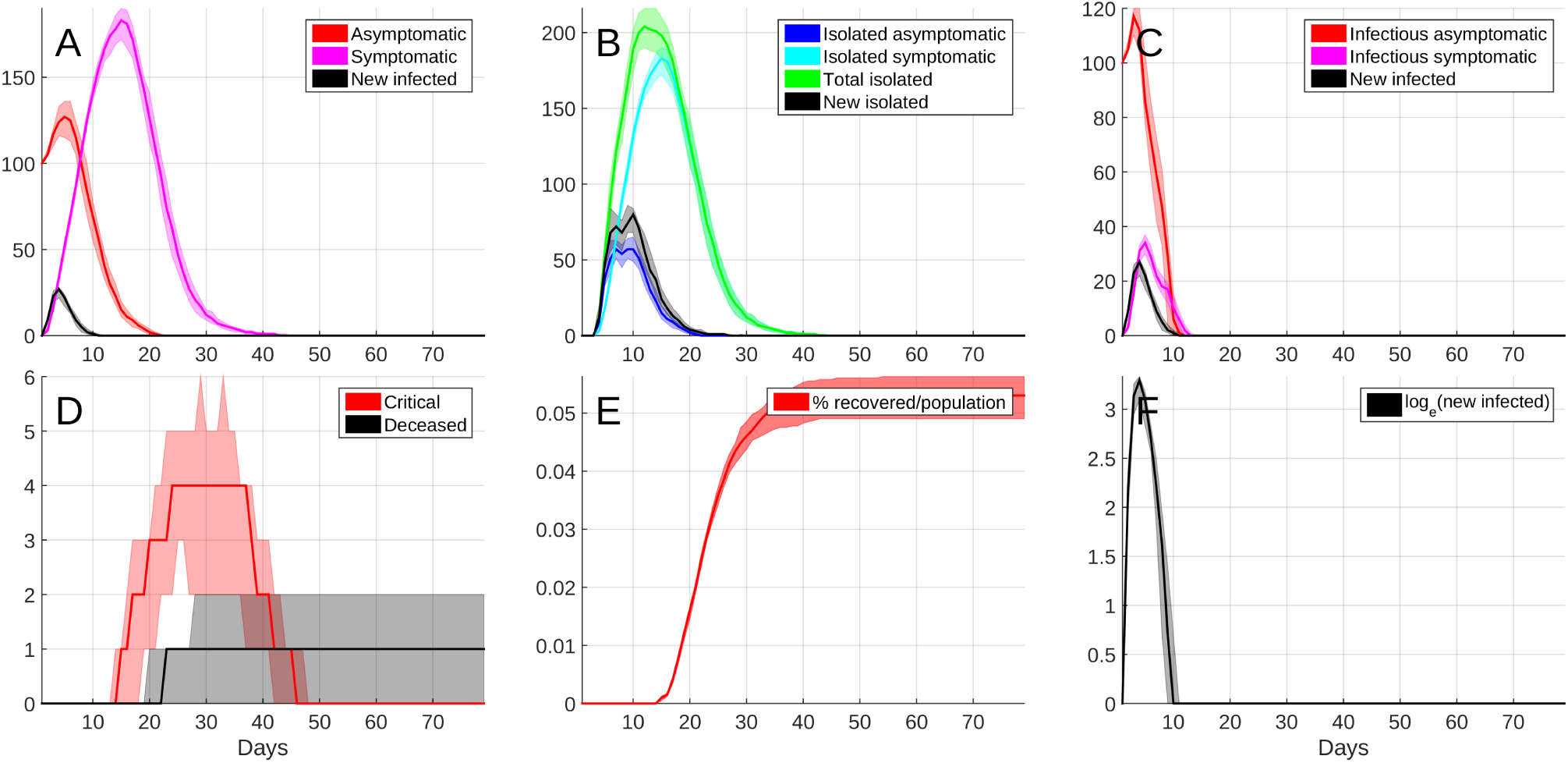
Example of slow-down solution obtained for *R*_0_ = 2, *δT_Q_ =* 2 days and *N_a_ =* 3. The coloured areas represent the envelopes of the 50% of models centred on the median model corresponding to the solid curves. The origin of the time axis corresponds to the beginning of the post-containment period.

**Critical solutions**. An example of a critical solution is shown in Figure 6. These solutions correspond to situations where the effective reproduction number ≈ 1. In this case, neither the exponential divergence not the damping are observed at long term. Instead, after a short period of exponential growth, the number of asymptomatic and symptomatic people oscillates around a constant level during a long duration (Fig. 6A). Similar oscillations are observed in the other curves. The instantaneous number of critical patients, *N_c_* is maintained at relative high levels (*N_c_* ≈ 50 − 100, Fig. 6D), and the cumulative number of deceased persons, ∑*N_d_* augments linearly. Contrarily to what is observed for the controlled slow-down solutions of Figure 5, the 50% envelopes of the critical solutions are very wide. This indicates that the stochastic process is very unstable due to the presence of a bifurcation between diverging and damped solutions (18, 19). The long-period oscillations visible in several curves (6A-D) may not be attributed to seasonal effects as described by Aron and Schwartz but could be due to stochastic resonance (20).

**Fig. 6.**
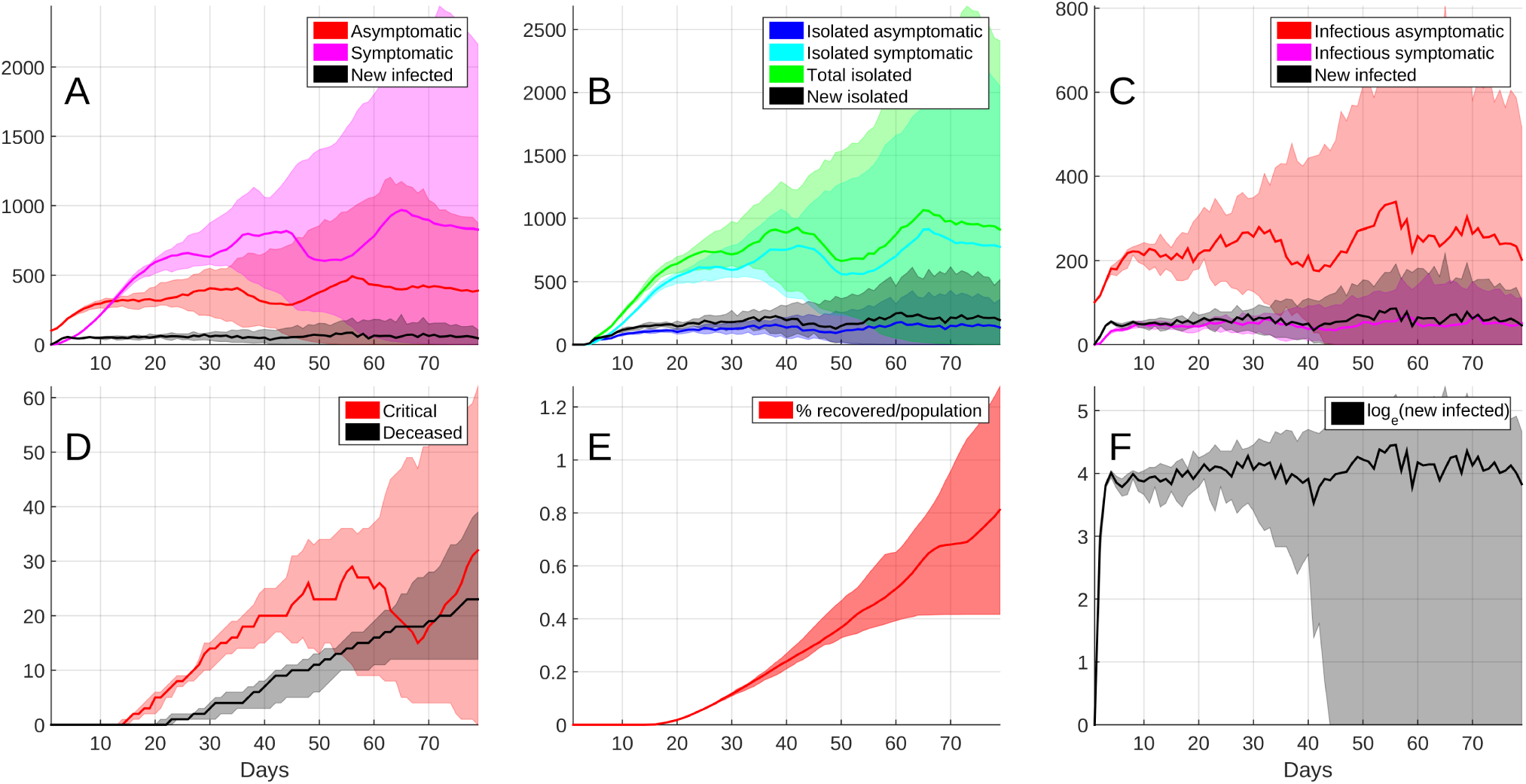
Example of critical solution obtained for *R*_0_ = 4, *δT_Q_* = 2 days and *N_a_* = 2.3. The origin of the time axis corresponds to the beginning of the post-containment period.

**Domains of solutions**. We performed a set of simulations in order to explore the solution domain (*R*_0_, δ*t_q_, N*_a_) in the limits given above. Figure 7 shows the sub-domains obtained for 4 values of *R*_0_ = 1.5,2.0,3.0,4.0. All simulations have been performed with an initial number of contagious persons *Z_I_* = 100. This number, although realistic, is probably too large, and the results shown in the Figures must be proportionally divided or multiplied to consider a different value for *Z_I_*.

**Fig. 7.**
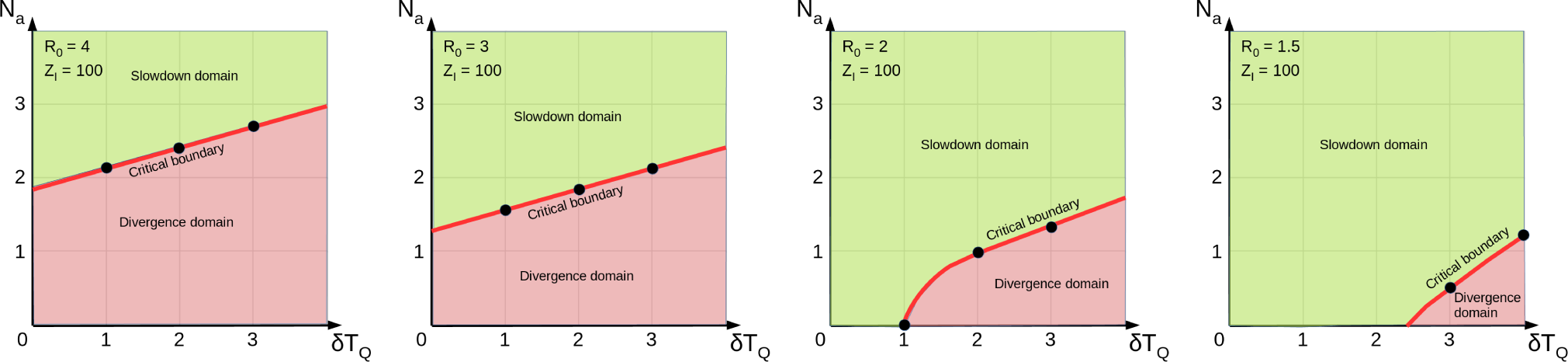
Solution domains (*δT_q_, N_a_*) obtained for *R*_0_ =4,3,2,1.5. *δT_Q_* is the time-delay elapsed between the apparition of symptoms in a patient and the moment when this patient is placed in quarantine. *N_a_* is the average number of asymptomatic persons detected and placed in quarantine when a symptomatic patient is detected. The green domains represent the models for which the epidemic spread is controlled. The red domains are the set of model for which the epidemic spread is uncontrolled with an exponential growing. The solid red lines represent the boundary between both domains and where the models are unstable with oscillations and large fluctuations.

The sub-domain for *R*_0_ = 4 corresponds to a situation where no social distancing is applied. This *R*_0_ value is slightly smaller than the value found before the beginning of the containment, i.e. when no social distancing policies were given, in the model shown in Figure 3. As can be observed in the corresponding graph of Figure 7, no slowing down of the epidemic spread can be obtained without both detecting symptomatic and asymptomatic people. In the most favourable case when δ*t_q_* = 1 day, at least 2.5 and even better 3 asymptomatic people must be detected each time a symptomatic patient is identified. Increasing *δt_Q_* worsens the situation with the necessity to detect a larger number of asymptomatic persons.

When social distancing is applied, we may expect a decrease of *R*_0_. In such a case, the smaller the *R*_0_ the wider the slowing down solution domain (Fig. 7). We observe that even a reduction of *R*_0_ from 4 to 3, an objective that seems very likely to reach, the slowing down can be obtained with seemingly reasonable conditions: δ*t_q_* = 2 days and *N_a_ >* 2. The solution domains for *R*_0_ = 2 and *R*_0_ = 1.5 offer solutions where it may be unnecessary to systematically detect symptomatic people to obtain a damping of the epidemic spread. However, it must be emphasised that these solutions do not converge quickly and they should only be considered as limit cases.

## Concluding remarks

The model used in the present study is an improved version of the stochastic model proposed by Allali et al.. The improvements concern the explicit distinction of asymptomatic and symptomatic people and the introduction of a log-normal distribution ζ*_a_/_s_* to represent the switching period between asymptomatic and symptomatic (Table 1). A non-uniform contagiousness function ξ*_c_* is represented by a gamma distribution (Table 1). This improved model allows to simulate the effects of several sanitary measures during the decontainment stage in Guadeloupe: i) the detection and isolation of symptomatic patients, ii) the testing and isolation of asymptomatic people, iii) a global reduction of the basic reproduction number through social distancing (21).

The simulations performed for different combinations of the three key parameters, *R*_0_, *δT_Q_* and *N_a_*, allowed to identify 3 main types of solutions: diverging exponential, slowdown damped and critical. These classes of solutions define two domains in the parameter space where conditions conduct either to an exponential spread of the disease or, instead, to a rapid damping of the exponential growth and a cease of the epidemic spread (Fig. 7). Critical solutions, where no exponential divergence occurs but where a sustained production of infected people remains, are located on a narrow boundary separating the two domains (red solid lines in Fig. 7).

Our simulations show that relatively wide domains of slowdown solutions exist provided the basic reproduction number *R*_0_ ≤ 2 (Fig. 7). These solutions are clearly those that should be engaged to control the epidemic spread. Owing to the fact that in normal conditions without any social distancing measures *R*_0_ ≈ 4.5 in Guadeloupe (1), it can be concluded that the condition *R*_0_ ≤ 2 can only be obtained by applying some social distancing measures with no strict containment excepted for all symptomatic patients and a subset of asymptomatic people.

At the time of completing the present paper, the French Government just issues his post-containment policy based on a mapping of the present (i.e. April 30, 2020) sanitary conditions in each French department. Guadeloupe belongs to the most favourable cases, with a sharp decrease of the number of new infected people (see Fig. 3A,C). This success certainly results from the conjunction of several positive factors, in particular: i) the fact that the epidemic spread started a few days before the beginning of the containment produced a rapid decrease of *R*_0_; ii) the island nature of Guadeloupe enabled a tight control of possibly infected incoming passengers; iii) an efficient and coordinated actions from all sanitary and administrative authorities resulting in clear and coherent messages sent to the population. This good control of the epidemic spread was already visible in the study by Allali et al. where all model solutions indicated a rapid decrease of *R*_0_. The management of the post-containment period strongly depends on the number *Z_I_* of infected people present in the territory at the beginning of the period. This number depends on the fraction of asymptomatic people who may concur to the propagation of the virus. It is commonly assumed that asymptomatic people represent about 20%, as used in our model. However, values up to 40% seem possible (11, 12). In the case of Guadeloupe, considering that 12 patients are deceased from COVID-19 by May 1, 2020, and taking an IFR = 0.7% (11), we find a total of 1700 infected persons, representing less than 0.4% of the population of Guadeloupe. Using these figures, at the beginning of the post-containment period scheduled on May 11, 2020, we may expect that the situation in Guadeloupe will correspond to a pre-epidemic situation, where only some tens of infected persons will be present on the territory among a non-immune population. In this case, one may envision that sanitary procedures successfully applied by several countries to contain and quickly stop the epidemic spread at its very beginning are likely to be successful in Guadeloupe. In the case of Singapore (22), spread control was mainly due to early detection of cases through testing and contact tracing around known cases in a way similar to our modelling assumptions described above. The case of Singapore shows that such procedures can be applied without creating a major disruption to daily living.

Returning to the case of Guadeloupe, we must consider that certainly not all symptomatic patients will be detected in the δ*T_Q_* delay and that social distancing will not be applied in several isolated groups of people. Consequently, a safety margin must be kept in mind when looking for solutions, and our simulations show that *R*_0_ = 2, δ*T_Q_* = 2 days and *N_a_* = 3 (Fig. 5) ensure an efficient slowing down with a very small number of critical and deceased people. This simulation also shows that, at the maximum of the crisis, about 80 symptomatic and asymptomatic people will have to be detected and isolated every day (“New isolated” curve in Fig. 5B). Supposing that only 10% of tested persons are really infected asymptomatic, it is then necessary to be able to perform at least 1000 tests per day to reach the objective of an average of 3 asymptomatic persons detected for 1 symptomatic. This objective corresponds to a slowdown solution in all four domains of Figure 7 with a preference for the cases of Fig. 7C,D to ensure a sufficiently large distance with the critical boundary. This clearly supposes the involvement of a numerous team of sanitary people to accomplish this task. In addition, the detection of clusters of contaminated people belonging to groups that do not apply social distancing rules should deserve a particular attention (12, 23).

## Data Availability

Data are available on the Santé Publique France website

## Supplementary Note 1: Model parameters

